# Real-World Effectiveness and Safety of Evolocumab in Very High-risk Atherosclerotic Cardiovascular Disease Patients with Acute Ischemic Stroke

**DOI:** 10.1101/2023.06.25.23291877

**Authors:** Ting Zhang, Yajing Zhang, Yun Yang, Haibing Liao, Xun Li, Ran Liu, Xueqing Liu, Liqin Yang, Wei Yue

**Author notes:** Corresponding author: Wei Yue, Department of Neurology, Tianjin Key Laboratory of Cerebrovascular and Neurodegenerative Diseases, Tianjin Huanhu Hospital, Clinical College of Neurology, Neurosurgery and Neurorehabilitation, Tianjin Medical University, Tianjin, China, 300350. E-mail address (Wei Yue). Ting Zhang and Yajing Zhang contributed equally to this work.

## Abstract

**Background:** We investigated evolocumab’s real-world effectiveness and safety on a background of statin therapy in the acute phase of ischemic stroke (IS) patients with a very high-risk of atherosclerotic cardiovascular disease (ASCVD).

**Methods:** A real-world, single-center, retrospective study was conducted in the neurology department at Tianjin Huanhu Hospital in China. Patients were divided into two groups: evolocumab treatment (140 mg every two weeks) or the standard of care (SOC) group. The primary efficacy outcome of the study was the achievement of a targeted lipid control rate and the incidence of major adverse cardiovascular events (MACE) by the end of the follow-up. Propensity score matching (PSM) analysis was utilized to account for confounding factors between groups. Survival analysises were performed using the Kaplan-Meier method.

**Results:** 1080 AIS patients with very high-risk ASCVD were recruited. After PSM, there were 528 individuals, with 206 in the evolocumab group and 322 in the standard of care (SOC) group. At 12 months of follow-up, the proportion of LDL-C < 1.4mmol/L and ≥50% reduction was 44.9% in the evolocumab group, compared with only 3.1% of SOC-treated patients (*p* = 0.000). The median follow-up time for clinical events was 15 months. The evolocumab group was associated with a lower risk of cerebrovascular events compared to the SOC group (HR, 0.45; 95% CI, 0.23-0.89; *p* = 0.022) but did not significantly reduce the incidence of cardiovascular events (HR, 0.51; 95% CI, 0.17-1.51; *p* = 0.224), event-related deaths (HR, 0.71; 95% CI, 0.34-1.46; *p* = 0.349), or MACE (HR, 0.71; 95% CI, 0.44-1.06; *p* = 0.089).

**Conclusions:** This real-world study suggested that evolocumab on a background of statin reduced the LDL-C levels significantly with a well-tolerated profile and lowered the incidence of recurrent cerebrovascular events in the very high-risk ASCVD patients with AIS in China.

## Introduction

Based on national stroke statistics in 2019, the age-standardized mortality rate of cerebrovascular disease in China was 149.49/100000, accounting for 22% of total deaths. Acute ischemic stroke (AIS) constituted about 80% of cerebrovascular disease. Due to the aging of the population, the prevalence of AIS in China showed an upward trend from 2005 to 2018, which caused a considerable disease burden to patients, families, and healthcare systems^1^. Although the causes of AIS are heterogeneous (e.g., cardioembolic and noncardioembolic that includes atherosclerosis, small vessel disease, and undetermined), the atherosclerotic disease was highly prevalent in 90% of the cases^2^. In patients with ischemic stroke, those with atherosclerosis have a much higher risk of major vascular events within 5 years than those without atherosclerosis^3^. The risk of atherosclerotic AIS is probably related to the disruption of a balance between instability (“activation”) and healing (“passivation”) of an atherosclerotic plaque^4^. Lipid-lowering treatments will help plaque healing and reduce the incidence, recurrence, and mortality in AIS with atherosclerotic evidence^5^.

Elevated low-density lipoprotein cholesterol (LDL-C) is a risk factor for atherosclerotic cardiovascular disease (ASCVD) events. Randomized controlled trials (RCTs) have demonstrated that lipid-lowering treatments are effective for primary and secondary AIS prevention^6^. The TST study indicated that AIS patients with a target LDL-C level of less than 70 mg/dl (1.8mmol/L) had a lower risk of subsequent ASCVD events than those with a target range of 90-110 mg/dl (2.31-2.83mmol/L)^7^. Recent data from RCT showed that when added to statin therapy, ezetimibe reduced the LDL-C level from 1.8 mmol/L to 1.4 mmol/L and provided an additional benefit of lowering the ASCVD risks (absolute risk difference, 2.0 percentage points; HR, 0.936; *p*=0.016)^8^. The 2018 American College of Cardiology (ACC)/American Heart Association (AHA) cholesterol Clinical practice Management guidelines further subdivided the high-risk ASCVD patients into very high-risk and not very high-risk based on the cardiovascular risk levels^9^. 2019 European Society of Cardiology (ESC) and the European Atherosclerosis Society (EAS) guidelines provided different risk-based LDL-C goals: secondary prevention for patients at very high-risk, an LDL-C reduction of ≥50% from baseline and an LDL-C goal of <1.4 mmol/L (55 mg/dL) were recommended (Class I, Level A)^10^. Also, a clinical guide by the Hellenic Stroke Organization and the Hellenic Atherosclerosis Society recommended that patients with ischemic stroke(IS) or transient ischemic attacks (TIA) should be treated with an LDL-C target<55 mg/dL (1.4 mmol/L) and at least 50% reduction of baseline LDL-C levels^11^. These guidelines have indicated that the lower the achieved LDL-C values, the lower the risk of future cardiovascular events.

Although statin treatment was the cornerstone of dyslipidemia management and had an established role in AIS prevention^12^, the real-world effectiveness of using high-intensity statin therapy in the Chinese population was still controversial. Many Chinese AIS patients had limited benefits from high-intensive statin treatment and failed to achieve the lipid treatment targets. The incidence of statin-related liver dysfunction and myopathy in Chinese patients treated with high-intensity statins was much higher than in European populations^13, 14^.

Proprotein convertase subtilisin/kexin type 9 (PCSK9) inhibitors are a new class of lipid-lowering drugs that can precisely target the disease pathway to enhance atherosclerotic plaque healing^15^. Several studies published since 2015 demonstrated that PCSK9 inhibitors (evolocumab) represented a promising approach for a rapid, profound reduction in LDL-C and reduced the risk of cardiovascular events, with a well-tolerable safety profile^16^. PCSK-9 inhibitors could be considered beneficial lipid-lowering drugs in addition to statins or ezetimibe. However, we have not seen data about the use of PCSK9 inhibitors in Chinese AIS patients.

Our study aimed to investigate evolocumab’s real-world effectiveness and safety in very high-risk ASCVD patients with AIS.

## Methods

### Study Design and Patients

This retrospective study was conducted at a single center between September 1st, 2019, and September 1st, 2021, in which AIS patients with a very high risk of ASCVD were recruited from the neurology department at Tianjin Huanhu Hospital. Clinical events were followed up until March 1st, 2023.

The primary efficacy outcome of the study was the incidence of major adverse cardiovascular events (MACE) by the end of the follow-up period. MACE was defined as a composite of various cardiovascular events, including acute coronary syndrome (ACS), myocardial infarction (MI), stable or unstable angina, coronary or other arterial revascularization, cerebrovascular events such as stroke or TIA, and event-related deaths. Another primary efficacy outcome was achieving a targeted lipid control rate, specifically LDL-C <1.4 mmol/l and a reduction of ≥50%, and non-HDL-C<2.2 mmol/L from baseline to 12 months of follow-up.

The other efficacy outcomes of the study included the percentage change of LDL-C, total cholesterol (TC), triglycerides (TG), high-density lipoprotein cholesterol (HDL-C), apolipoprotein A1 (ApoA1), and apolipoprotein B (ApoB) from baseline to 12 months of follow-up. Additionally, the incidence of cardiovascular events such as ACS, MI, stable or unstable angina, coronary or other arterial revascularization, cerebrovascular events such as stroke or TIA, and event-related deaths at the end of follow-up were also assessed separately.

The primary safety outcomes of the study were the incidence of adverse events (AEs) and serious adverse events (SAEs). In this study, adverse reactions with a frequency of ≥1% in the drug insert of evolocumab, which were higher than those in the placebo treatment group, were selected as the primary observation endpoint for safety outcomes.

Patients who met the following criteria are eligible for the study: 1) aged 18 to 90 years; 2) hospitalized within 14 days of AIS onset; 3) atherosclerotic origin; 4) very high-risk of ASCVD and receive lipid-lowering therapy, either evolocumab or standard of care (SOC) treatment. Patients were ruled out if they had: 1) coexistence of intracranial hemorrhagic disease; 2) individuals exhibiting indisputable evidence of cardiogenic embolism; 3) patients treated with PCSK-9 inhibitors before; 4) a modified Rankin Scale (mRS) score before hospitalization of more than 2. The Institutional Ethical Committee of the Tianjin Huanhu Hospital approved the study protocol with an exception to the requirement of written informed consent.

Very high-risk ASCVD was defined as patients with more than two serious ASCVD events or one serious ASCVD event with more than two high-risk factors recommended by the Chinese expert consensus on lipid management. Clinical ASCVD events included ACS, MI, stable or unstable angina, coronary or another arterial revascularization, stroke, TIA, or peripheral artery disease (PAD), including aortic aneurysm. High-risk factors included poly-vascular disease (2-3 arterial lesions with ischemic symptoms in a coronary artery, cerebral artery, or peripheral artery); premature coronary heart disease (male <55 years old, female <65 years old); familial hypercholesterolemia or baseline LDL-C > 4.9mmol/L; previous history of coronary artery bypass grafting or percutaneous coronary intervention; diabetes mellitus (DM); hypertension; chronic kidney disease (stage 3/4); smoking; LDL-C ≥2.6mmol/L after a maximum tolerable dose of statins^17^. AIS was defined by World Health Organization criteria as a sudden focal neurologic deficit persisting longer than 24 hours and confirmed by brain CT or MRI. We define atherosclerotic stroke as IS patients of presumably atherosclerotic origin. The patients had evidence of atherosclerosis: including intracranial, carotid, vertebral, and aortic arch atherosclerotic plaques/stenosis confirmed by vascular ultrasound, MRA, CTA, and angiography. SOC treatment is the standard lipid-lowering treatment. All lipid-lowering products available in Tianjin, China, were evaluated for prescription use (atorvastatin, fluvastatin, rosuvastatin, and Xuezhikang Capsule, which is an extract of Chinese red yeast rice containing a family of naturally occurring statins^18^).

### Data collection

The patient’s baseline and follow-up information were extracted from a stroke database established at our hospital by trained research staff. The collected data included patients’ demographics, medical history, laboratory, stroke severity, treatment, etc. The severity of stroke was measured using the National Institutes of Health Stroke Scale (NIHSS) score at admission and divided into three levels (NIHSS≤8; 9≤NIHSS≤15; NIHSS ≥16).

Blood samples were obtained in the early morning of the day after admission. An immunoturbidimetric assay measured all routine lipids and lipoprotein levels in serum samples (Beckman Coulter AU5800 automatic biochemical analyzer).

Patients with AIS received evolocumab 140 mg every two weeks combined with statins or SOC treatment, based on the physician’s suggestion and the choice of patients or their families. Patients are subjected to follow-up via either telephone or in-person consultations at the outpatient clinic.

### Statistics analysis

Continuous variables were presented as mean ± SD (95% CI) or median (interquartile range). Categorical variables were presented as numbers and percentages. The differences of continuous variables were compared using nonparametric tests for non-normally distributed data or T-tests for normally distributed data. The differences between categorical variables were compared using the X^2^ test or Kruskal – Wallis test, as appropriate. The propensity score-matched (PSM) method was applied to balance the confounding factors between the two groups. The selected variables to be potential confounders associated with clinical outcomes were age, gender, past medical history, etc. The evolocumab and SOC groups were paired at 1:2 using nearest matching with a caliper size 0.05. Survival analysis using Kaplan-Meier methods associated with log-rank tests was applied for MACE (cardiovascular events, cerebrovascular events, and event-related deaths). We performed subgroup analysis on the results of survival analysis to investigate the consistency of treatment effect across diverse patient subgroups and calculated the P value for interaction. Hazard Ratios (HRs) and 95% CIs were provided, with a corresponding P-value. All statistical tests are two-tailed, and statistical significance is set at *p < 0*.05, which were performed using SPSS 26.0 (SPSS, Chicago, IL) and R version 4.2.2.

## Results

### Baseline characteristics

Initially, the study involved registering 1080 patients with AIS who were at a very high-risk of ASCVD between September 1st, 2019, and September 1st, 2021. Following registration, a meticulous screening process was conducted, whereby patients with cardioembolism, intracranial hemorrhage, malignant tumors, mRS>2 before hospitalization, incomplete baseline information, and insufficient follow-up, etc., were excluded. Consequently, out of the initial 1080 registrants, 898 patients were deemed eligible and subsequently assigned to either the evolocumab or the SOC group (Figure 1).

**Figure 1.**
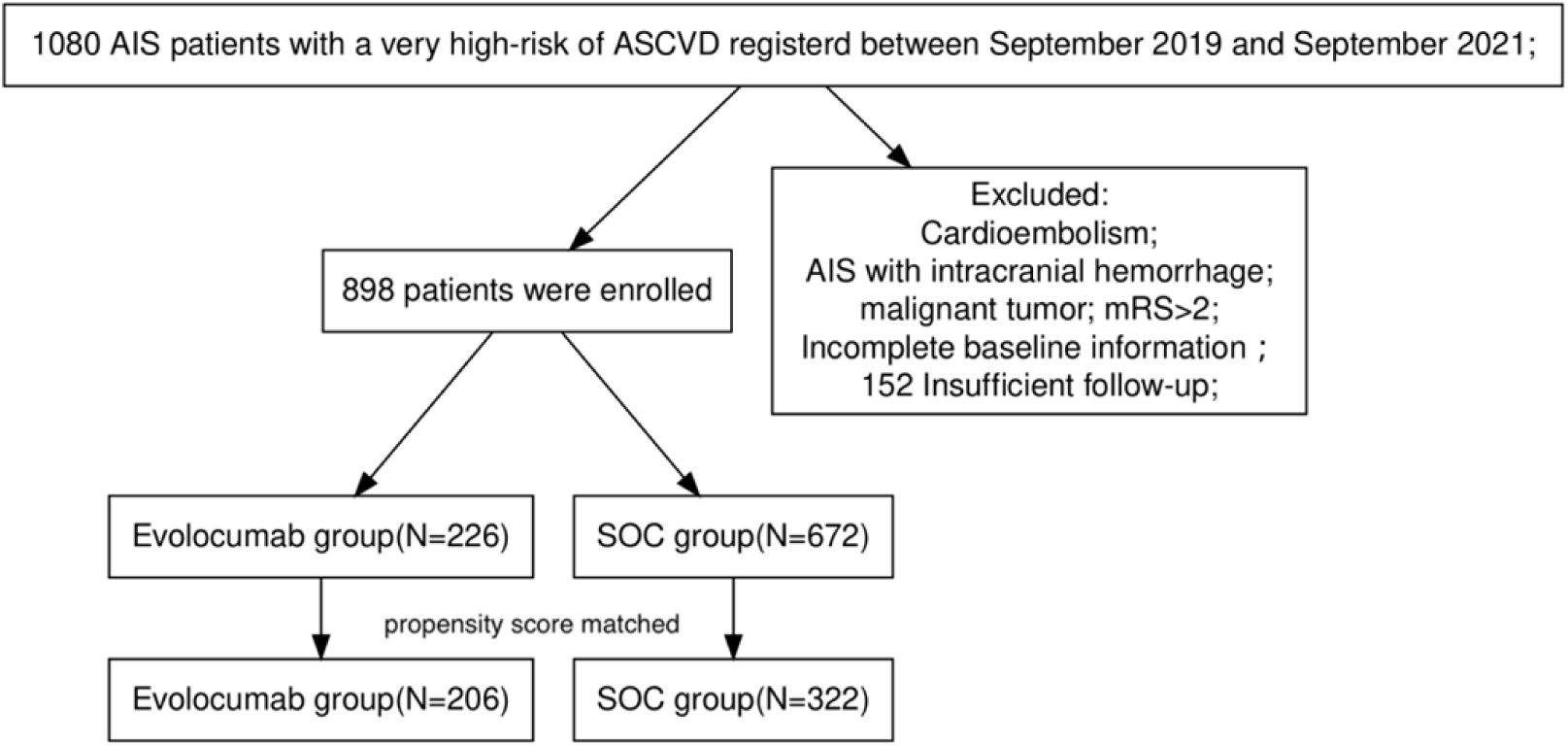
Patient Flowchart. ASCVD: atherosclerotic cardiovascular disease; AIS: acute ischemic stroke; SOC: standard of care;

Of 898 patients, 226 received evolocumab combined with statin therapy background, and 672 received SOC treatment. At baseline, only a minority of patients (14.1% vs. 16.1%) were treated with statins before hospitalization. The evolocumab group had a slightly lower median age (62.0 years, interquartile range, 55.0 to 69.0) compared to the SOC group (64.0 years, interquartile range, 56.0 to 71.0) (*p* =0.047) and a higher proportion of current smokers (51.8% vs. 43.3%, *p* =0.033). The evolocumab group also had a higher prevalence of diabetes (39.8% vs. 32%, *p* =0.043) and a lower prevalence of a family history (27% vs. 19%, *p* =0.015) compared to the SOC group. Notably, patients in the evolocumab group had a significantly higher NIHSS score at baseline compared to the SOC group (*p* <0.001). The other baseline characteristics, including gender, hypertension, coronary heart disease (CHD), stroke, and drinking, did not differ significantly between the two groups. These findings suggest that the evolocumab group had a slightly different baseline profile compared to the SOC group, with higher stroke severity and prevalence of diabetes and smoking. The patients’ TG, TC, LDL-C, and ApoB levels were higher in the evolocumab group than in the SOC group. Abnormal liver function (GGT and/or ALT) at baseline was more frequent in the evolocumab group. In order to mitigate the potential influence of confounding factors, PSM was used. As a result, the evolocumab Group was reduced to 206 patients, while the SOC Group was reduced to 322 patients, thereby ensuring comparability between the groups. Baseline characteristics of the two groups before and after PSM are presented in Table 1 and 2.

**Table 1.**
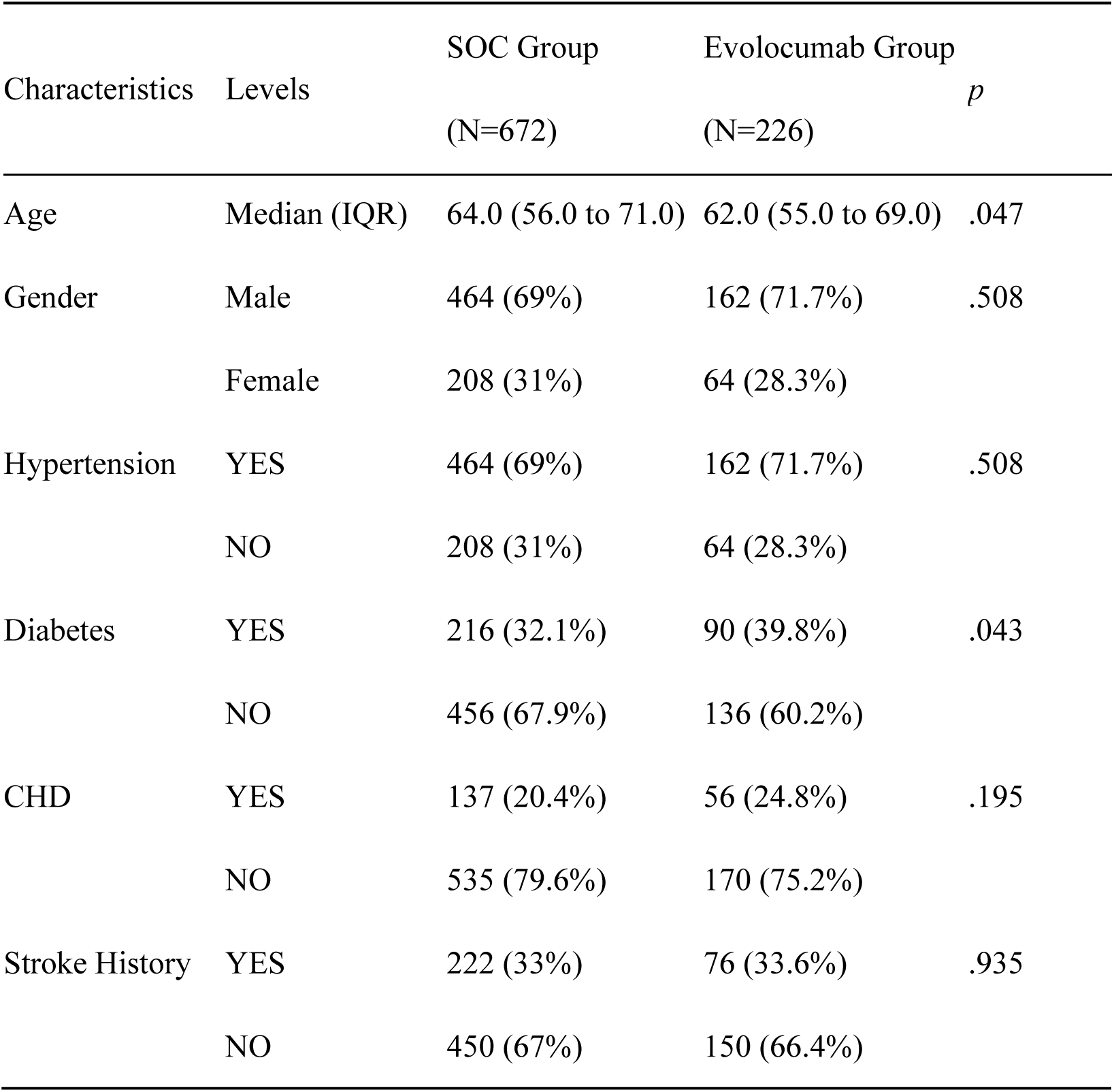
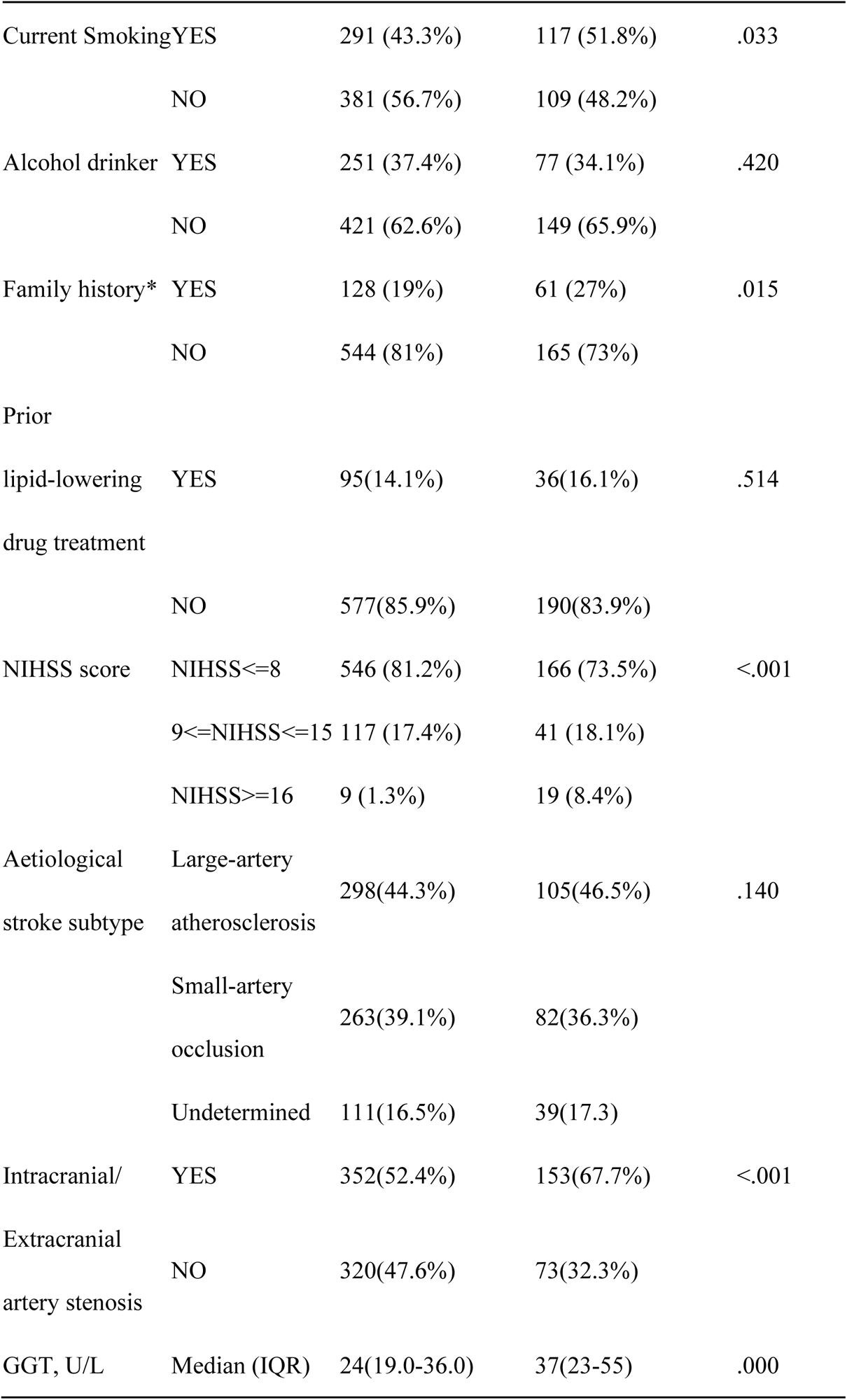
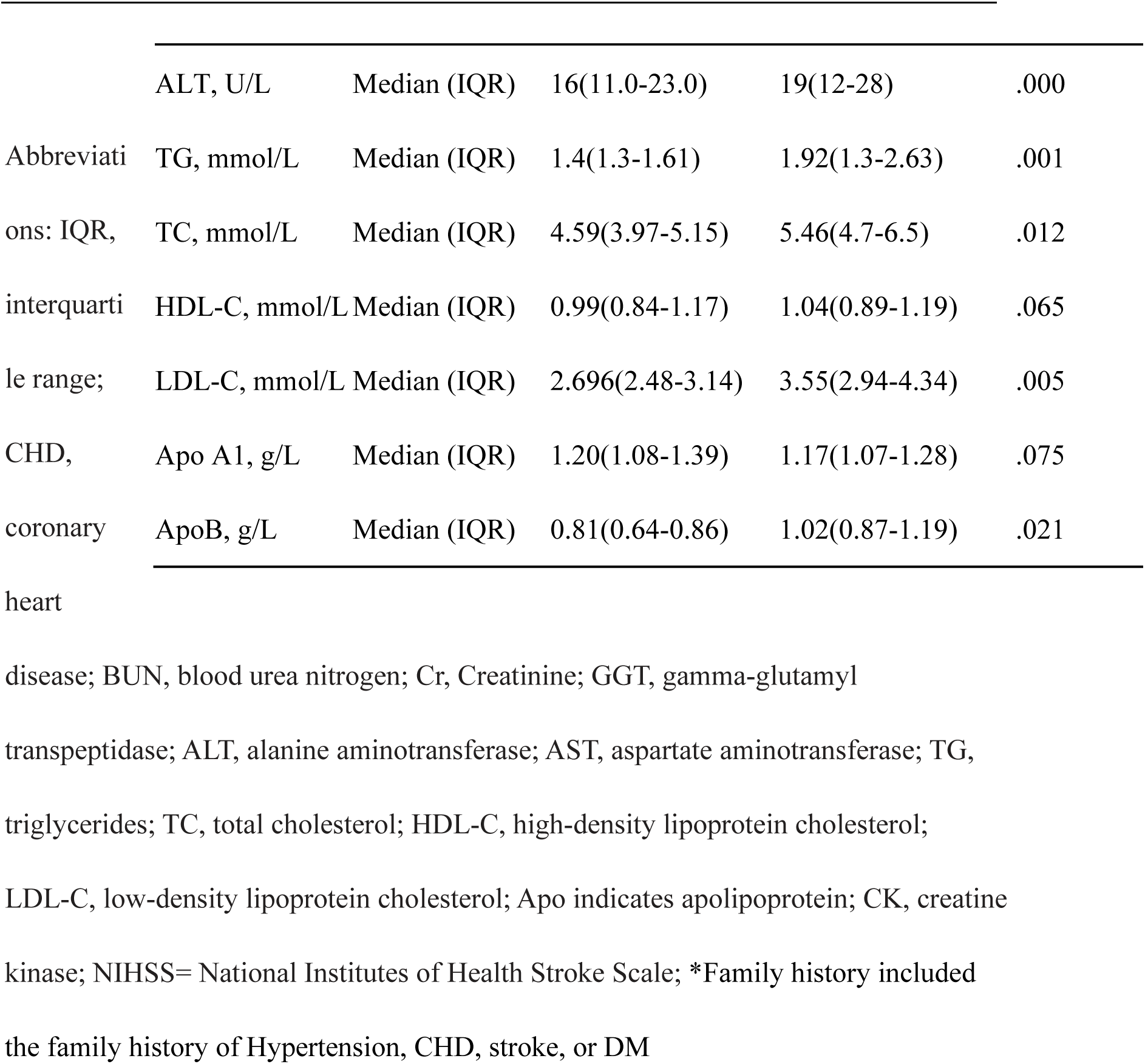
Demographic and characteristics of AIS patients before matching.

**Table 2.**
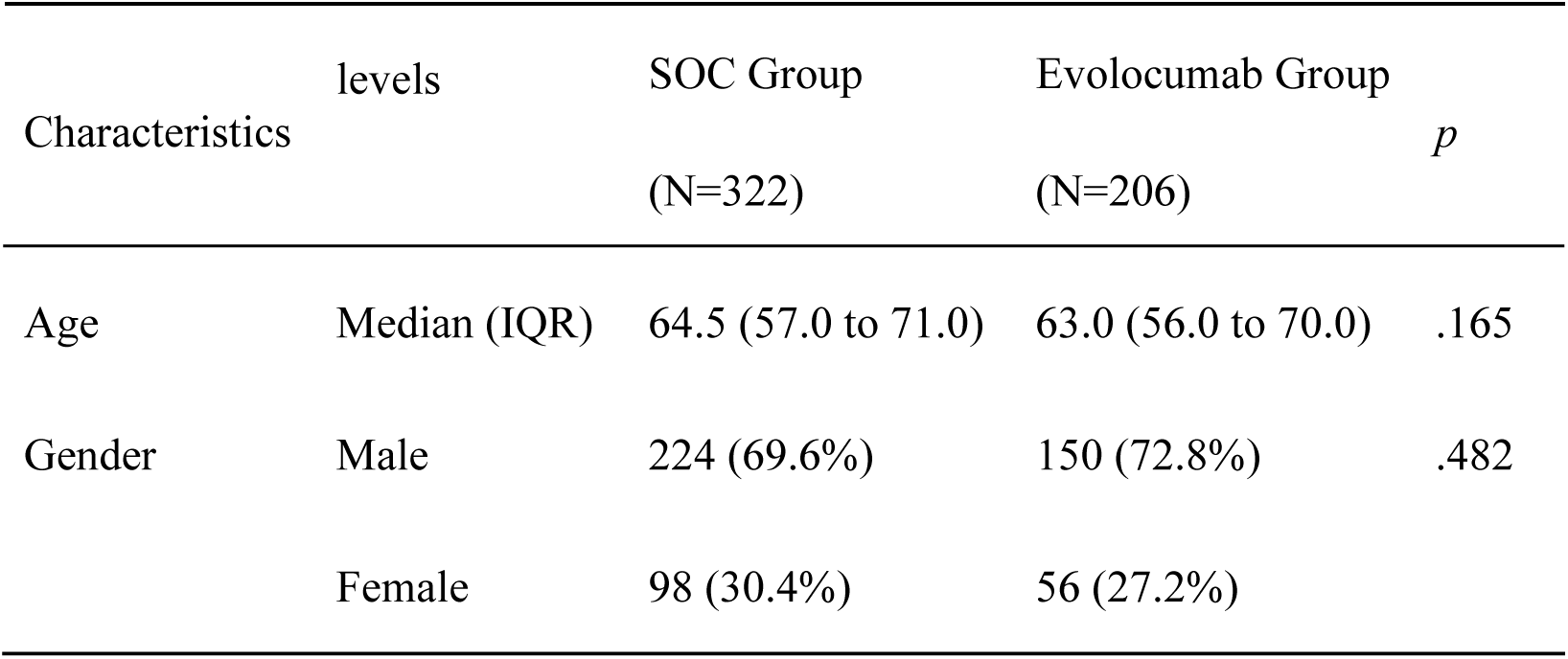
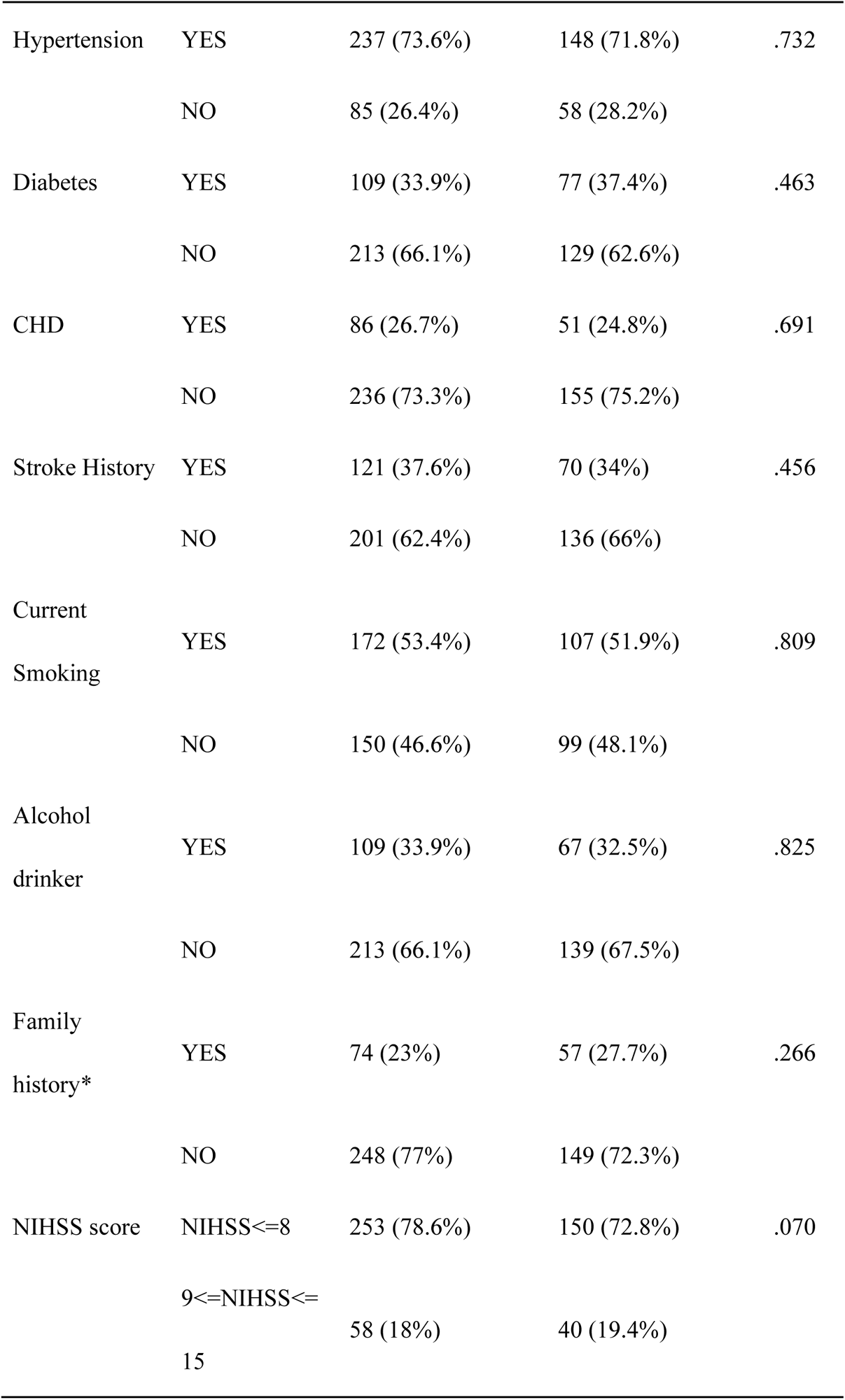
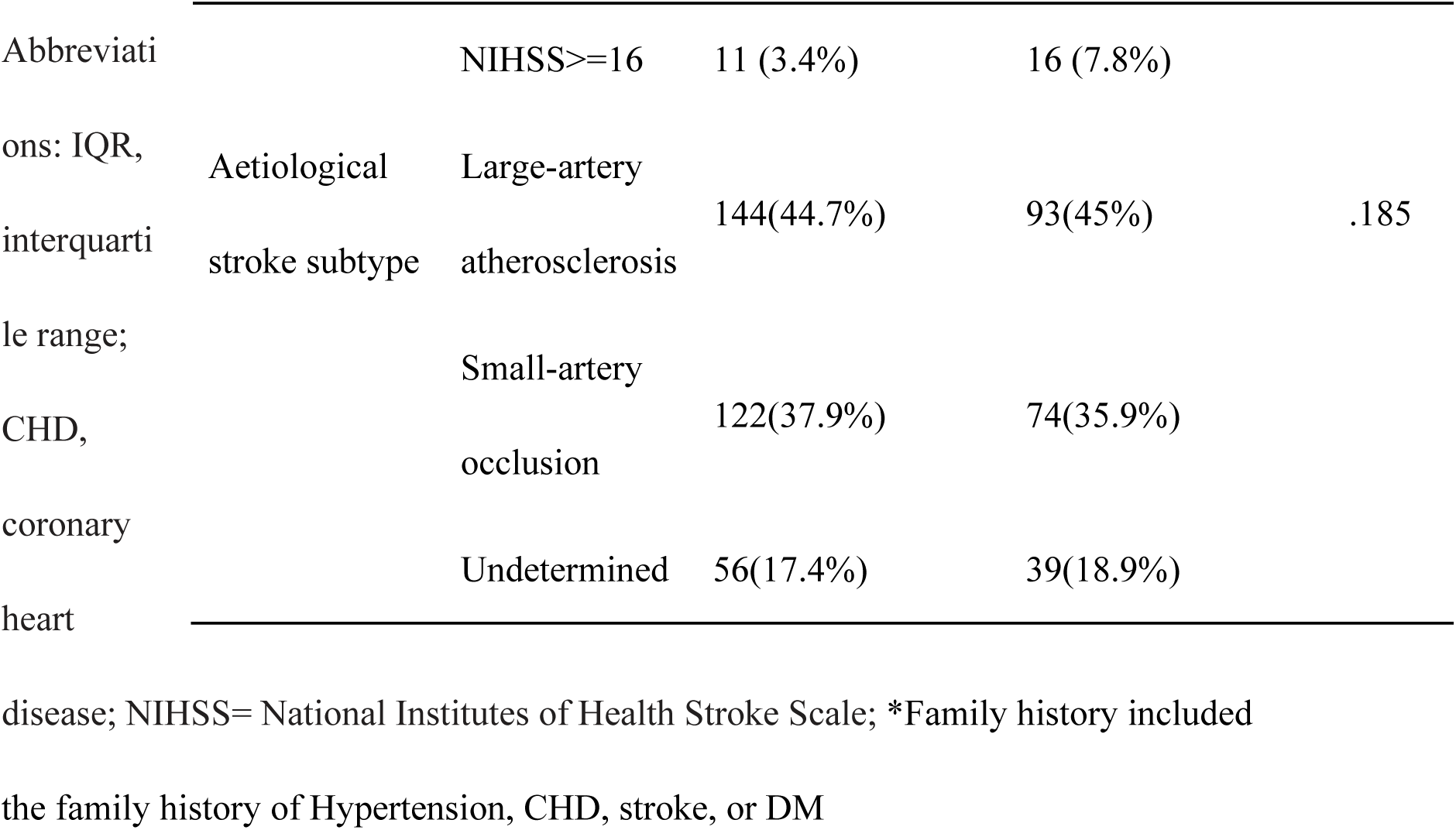
Demographic and characteristics of AIS patients after matching.

| Characteristics | levels | SOC Group<br>(N=322) | Evolocumab Group<br>(N=206) | *p* |
| --- | --- | --- | --- | --- |
| Age | Median (IQR) | 64.5 (57.0 to 71.0) | 63.0 (56.0 to 70.0) | .165 |
| Gender | Male | 224 (69.6%) | 150 (72.8%) | .482 |
|  | Female | 98 (30.4%) | 56 (27.2%) |  |
| Hypertension | YES | 237 (73.6%) | 148 (71.8%) | .732 |
|  | NO | 85 (26.4%) | 58 (28.2%) |  |
| Diabetes | YES | 109 (33.9%) | 77 (37.4%) | .463 |
|  | NO | 213 (66.1%) | 129 (62.6%) |  |
| CHD | YES | 86 (26.7%) | 51 (24.8%) | .691 |
|  | NO | 236 (73.3%) | 155 (75.2%) |  |
| Stroke History | YES | 121 (37.6%) | 70 (34%) | .456 |
|  | NO | 201 (62.4%) | 136 (66%) |  |
| Current<br>Smoking | YES | 172 (53.4%) | 107 (51.9%) | .809 |
|  | NO | 150 (46.6%) | 99 (48.1%) |  |
| Alcohol<br>drinker | YES | 109 (33.9%) | 67 (32.5%) | .825 |
|  | NO | 213 (66.1%) | 139 (67.5%) |  |
| Family<br>history* | YES | 74 (23%) | 57 (27.7%) | .266 |
|  | NO | 248 (77%) | 149 (72.3%) |  |
| NIHSS score | NIHSS<=8 | 253 (78.6%) | 150 (72.8%) | .070 |
|  | 9<=NIHSS<=15 | 58 (18%) | 40 (19.4%) |  |

|  |  |  |  |  |  |
| --- | --- | --- | --- | --- | --- |
| Abbreviations: IQR, interquartile range; CHD, coronary heart | | NIHSS≥16 | 11 (3.4%) | 16 (7.8%) | |
|  | Aetiological stroke subtype | Large-artery atherosclerosis | 144(44.7%) | 93(45%) | .185 |
|  |  | Small-artery occlusion | 122(37.9%) | 74(35.9%) |  |
|  |  | Undetermined | 56(17.4%) | 39(18.9%) |  |

disease; NIHSS= National Institutes of Health Stroke Scale; \*Family history included the family history of Hypertension, CHD, stroke, or DM

### Lipid Data

After 12 months of follow-up, the percentage change in LDL-C level was 59.96% (interquartile range, 45.38-69.1) in the evolocumab group (from a median of 3.65 mmol/L to 1.49 mmol/L) vs. 24.2% (interquartile range, 14.52-36.39) in the SOC group (from a median 2.66 mmol/L to 1.96 mmol/L), *p*=0.000. The changes in other lipid tests were significantly reduced in the evolocumab group compared to the SOC group, which reduced non-HDL-C levels by 58.14%, TG levels by 20.59%, TC levels by 49.62%, and ApoB levels by 53.27%. The HDL-C and Apo A1 were also significantly reduced compared to the baseline (*p* < 0.05 for all comparisons of evolocumab vs. SOC group). The proportion of LDL-C < 1.4mmol/L and ≥50% reduction was 44.7% in the evolocumab group compared with only 3.1% of SOC-treated patients (*p*=0.000). The percentage of participants achieving non-HDL-C < 2.2 mmol/L was 64.6% in the evolocumab group compared with 38.8% in the SOC group, *p*=0.000 (Table 3 and Supplementary materials).

**Table 3.**
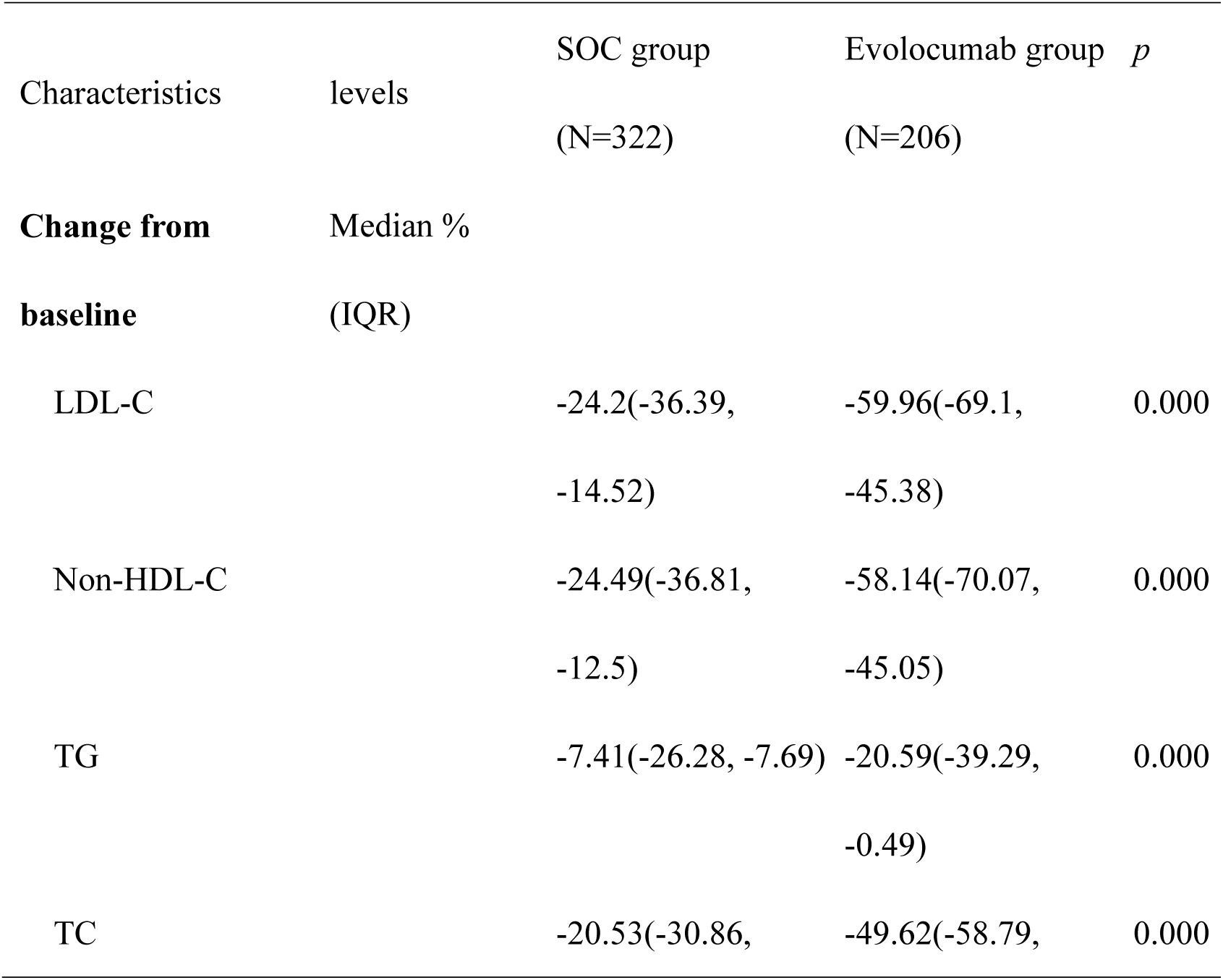
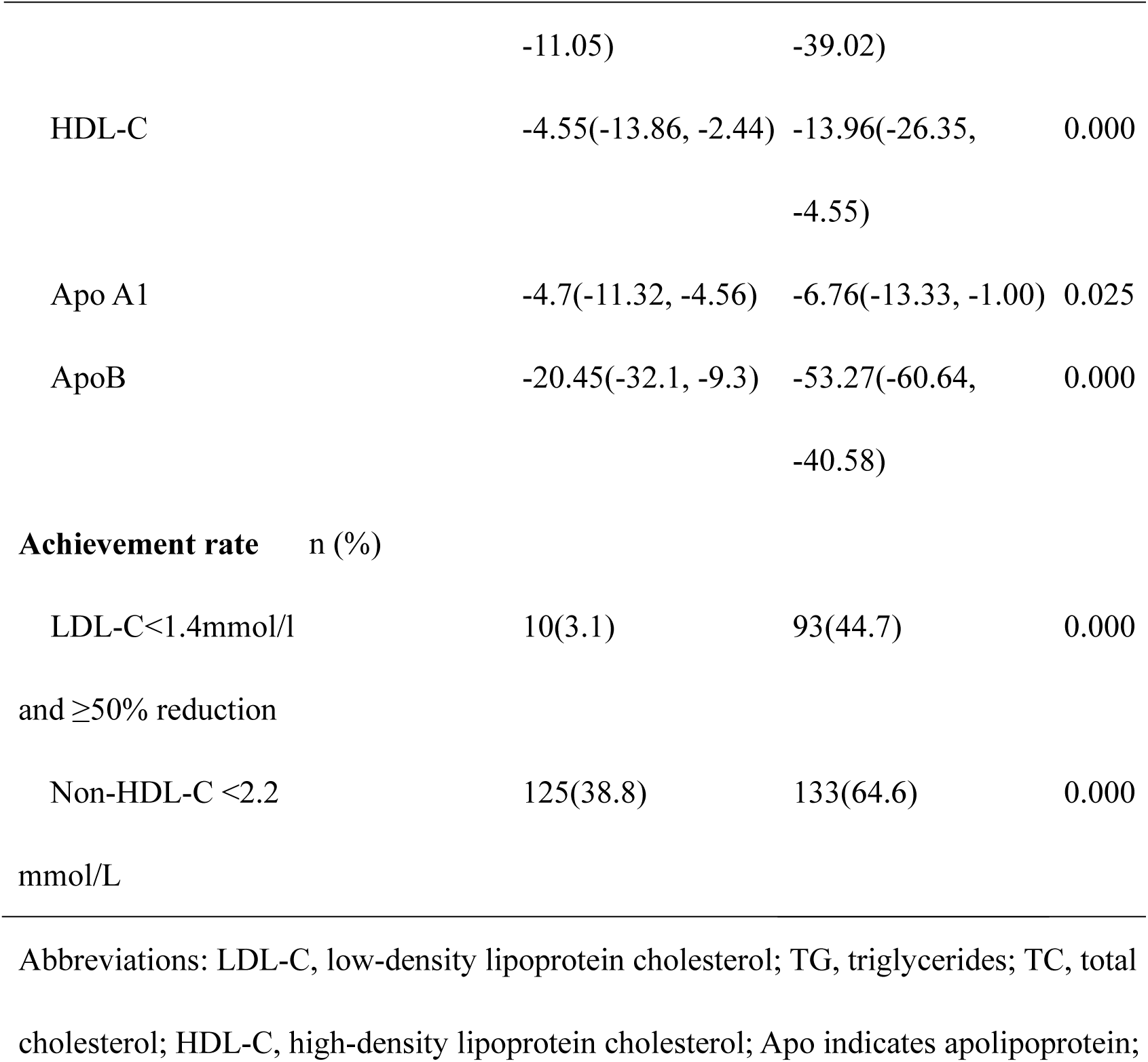
Changes in lipid levels between baseline and follow-up.

### Survival Analysis of the Clinical Events

During a median follow-up period of 15 months, survival analysis elucidated that the adjunctive use of evolocumab with statin therapy did not confer a significant reduction in the incidence of MACE (HR, 0.68; 95% CI, 0.44-1.06; *p* = 0.089), cardiovascular events (HR, 0.51; 95% CI, 0.17-1.51; *p* = 0.224), or event-related deaths (HR, 0.50; 95% CI, 0.34-1.46; *p* = 0.349). However, it is noteworthy that the intervention of evolocumab was significantly associated with a lower risk of cerebrovascular events (IS/TIA) when compared to the use of statin therapy alone (HR, 0.45; 95% CI, 0.23-0.89; *p* = 0.022). The Kaplan-Meier method demonstrated that the cumulative recurrence rate of cerebrovascular events at 12 months was 4% (95% CI, 1-7%) in the evolocumab group, compared to 7% (95% CI, 4-9%) in the SOC group. At 24 months, the recurrence rate was 8% (95% CI, 3-13%) in the evolocumab group, compared to 17% (95% CI, 9-25%) in the SOC group, P values were calculated with the use of log-rank tests (Table 4, Figure 2).

**Figure 2.**
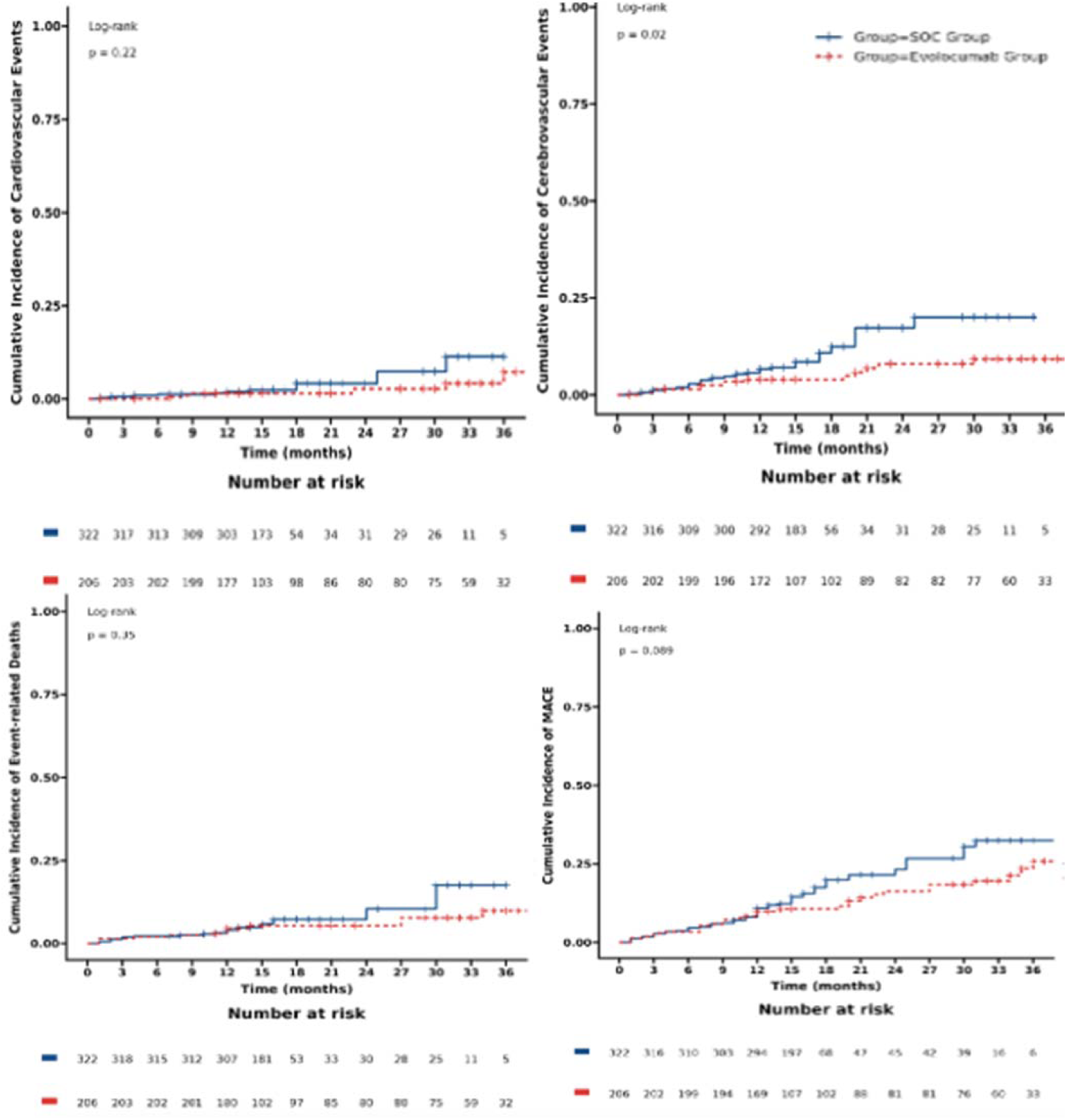
Cumulative Incidence of Clinical Events.

**Table 4.**
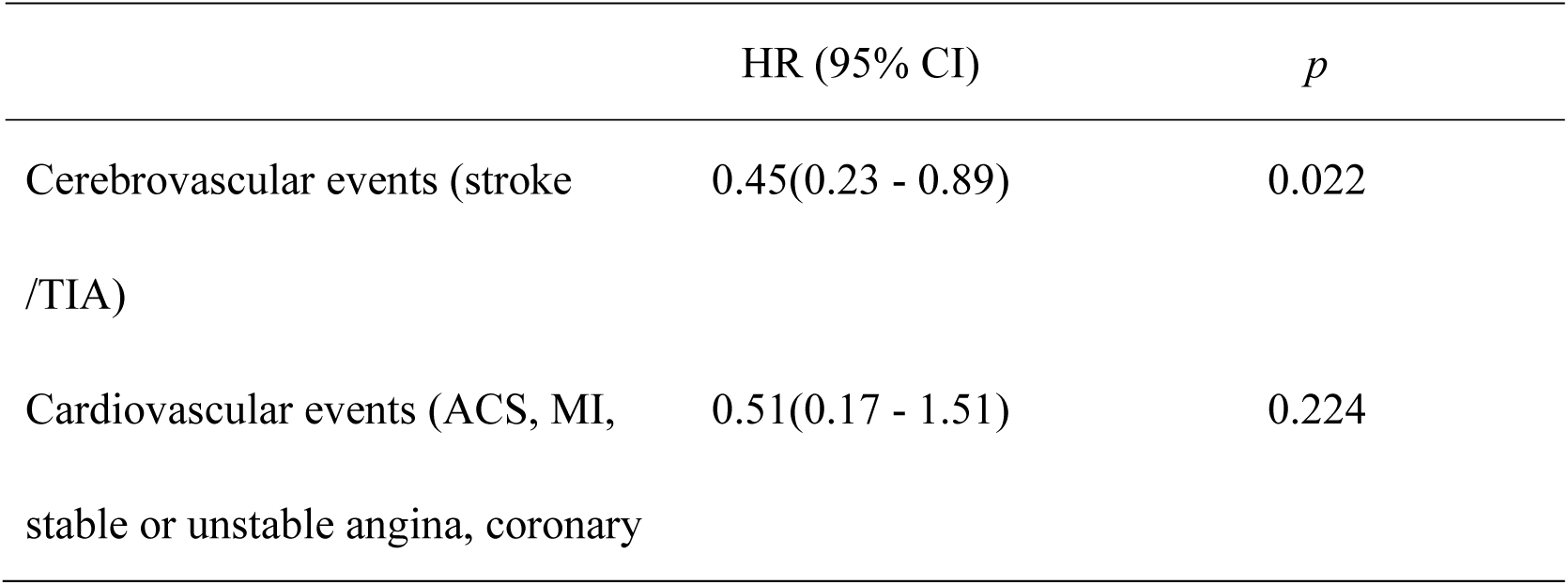
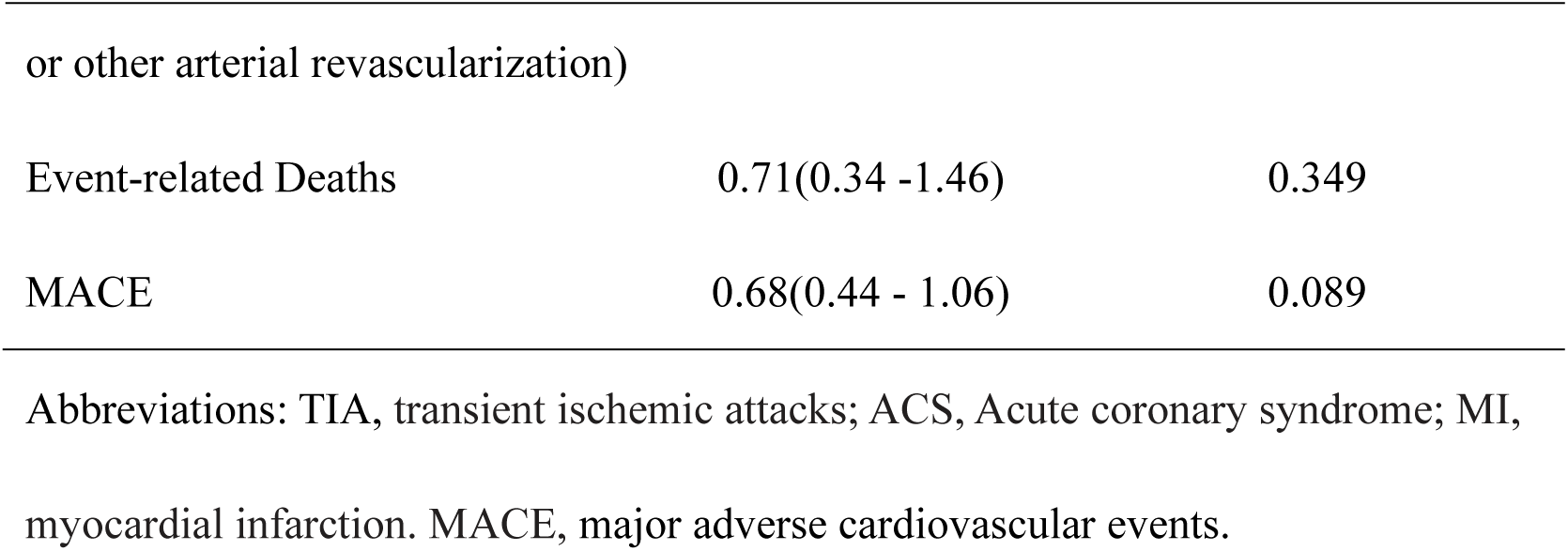
The Survival Analysis for the clinical endpoint events.

|  | HR (95% CI) | *p* |
| --- | --- | --- |
| Cerebrovascular events (stroke<br>/TIA) | 0.45(0.23 - 0.89) | 0.022 |
| Cardiovascular events (ACS, MI,<br>stable or unstable angina, coronary | 0.51(0.17 - 1.51) | 0.224 |
| or other arterial revascularization) |  |  |
| Event-related Deaths | 0.71(0.34 -1.46) | 0.349 |
| MACE | 0.68(0.44 - 1.06) | 0.089 |

Abbreviations: TIA, transient ischemic attacks; ACS, Acute coronary syndrome; MI, myocardial infarction. MACE, major adverse cardiovascular events.

In the subgroup analysis of survival analysis for cerebrovascular events, the efficacy of evolocumab was notably augmented in the subgroup of male patients, aged over 60 years, who had diabetes or CHD, smoked, manifested higher NIHSS scores, and exhibited evidence of intracranial/extracranial arterial stenosis (*p* < 0.050). Regarding the subgroup analysis of survival analysis for cardiovascular events, evolocumab’s benefit was more marked among patients who presented evidence of intracranial/extracranial arterial stenosis (HR, 0.13; 95% CI, 0.02-0.69; *p* =0.017). Similarly, the subgroup analysis for MACE demonstrated that the subgroup with intracranial/extracranial arterial stenosis also profited from evolocumab (HR, 0.59; 95% CI, 0.35-1.00; *p* = 0.050). No statistically significant subgroups were detected in the survival analysis for fatal events. We also performed a comparative analysis of the incidence rates of hemorrhagic stroke in both groups. No statistically significant differences were observed between the groups (HR, 0.94; 95% CI, 0.59-1.53; *p* = 0.621) (Supplementary materials).

Regarding safety outcomes, no substantial differences were observed between the evolocumab and SOC groups regarding the incidence of allergic reactions, neurocognitive events, musculoskeletal pain, and new-onset diabetes (Supplementary materials).

## Discussion

As a new class of cholesterol-lowering drugs, several studies have been published to demonstrate the clinical efficacy and safety of PCSK9 inhibitors (evolocumab) in patients with familial hypercholesterolemia^19^, stable ASCVD^20^, type 2 diabetes^21^ or ACS with hypercholesterolemia^16^. A prespecified analysis of cerebrovascular events in the FOURIER trial showed that evolocumab added to statins in patients with established atherosclerosis reduced ischemic cerebrovascular events with no increase in hemorrhage stroke in IS patients^22^. To the best of our knowledge, no previous study has reported evolocumab’s real-world effectiveness and safety on a statin therapy background in Chinese AIS individuals with very high-risk of ASCVD.

This study showed that evolocumab 140 mg once every two weeks on a background of statin therapy resulted in a significant reduction in LDL-C compared with statins alone among the AIS patients. Our analysis revealed that this therapeutic combination was linked with a reduced likelihood of cerebrovascular incidents, particularly in individuals with multiple risk factors (such as male gender, advanced age exceeding 60 years, diabetes or CHD, smoking, and higher NIHSS scores). In addition, patients with evidence of intracranial/ extracranial arterial stenosis experienced a lower incidence of recurrent cardiovascular and cerebrovascular events when treated with evolocumab. Also, evolocumab was well tolerated during the follow-up period and no significant difference was observed in AEs between the two groups.

Compared with previous clinical studies of other countries^21^, the LDL-C level at baseline was much higher in our study (3.0mmol/L vs. 2.38 - 2.81mmol/L). Since only 14.9% of them were treated by lipid-lowering drugs before admission. Although our study included urban individuals living in a relatively affluent city of China, the very high-risk ASCVD patients still lacked adequate awareness about lipid management. Based on The DYSISChina study, the LDL-C achieved rate among ASCVD patients was only 6.8%^23^, highlighting the importance of publicity and education on lipid management at the community and hospital level. We can strengthen communication between physicians and patients by providing advice on evidence-based education (such as distributing printed brochures and motivating physicians to complete survey forms), bridging the gap between guidelines and daily clinical practice^24^.

The LDL-C treatment difference in our study is about 35.76% (evolocumab minus placebo), which is close to the result of EVOPACS (40.7%)^16^ but is lower than FOURIER (59%)^20^ and BANTING (53.1%)^21^. The reasons underlying this gap are likely to be multifactorial. First, in the SOC group, more patients may be treated with high-intensity statins. Doctors are more likely to choose moderate-intensity statins combined with evolocumab due to this group’s high proportion of abnormal liver function. Second, a subgroup study of FOURIER found that evolocumab reduced the percentage of LDL-C for patients with lower baseline LDL-C levels more significantly^25^.

In real-world utilization, doctors recommend evolocumab to patients with baseline liver dysfunction who are more likely to be unable to tolerate high-intensity statins; and patients with higher baseline LDL-C who were not expected to reach the recommended treatment targets even if using high-intensity statin treatment. A scientific statement entitled ‘Safety of Statins and Related Adverse Events’ issued by AHA estimated that myopathy (unexplained muscle pain or weakness accompanied by CK increase > 10 times the ULN) occurred in < 1/1000 patients treated with maximum recommended doses and with even lower risk at lower doses. Severe hepatotoxicity is rare (about 1/10000). Dose-dependent asymptomatic elevations in transaminases above three times the ULN occurred in 1/100 people in clinical trials. But these are usually transient and typically not associated with signs or symptoms of liver disease^26^. However, statin pharmacokinetics is different in Asian and Western populations. The elevation rate of liver enzymes (> 3 times the ULN) and the risk of myopathy in Chinese patients were ten times higher than in European patients^13^. In addition, the China intensive lipid lowering with statins in acute coronary syndrome (CHILLAS) study showed that LDL-C levels declined by 20.2% in the moderate-dose statin group and 26.6% in the high-intensive statin group after three months follow-up period. The incremental LDL-C reduction was 6.4% but without clinically significant differences among the two groups and more safety events related to high-dose statin^14^.

Our finding suggests that evolocumab, in combination with statin therapy, may confer a protective effect against cerebrovascular events, although caution should be exercised when interpreting these results due to the limitations of Kaplan-Meier analysis. Initially, the cumulative recurrence curves of cerebrovascular events exhibited similarity for the first half-year; however, as time progressed, subtle discrepancies emerged among the curves representing the distinct treatment cohorts. In FOURIER, a comparable trend was noted, where the extent of the decline in the risk of clinical events escalated gradually with the passage of time, signifying that the conversion of reducing LDL-C levels into vascular clinical advantages necessitates a certain duration^20^.

Intracranial/extracranial arterial stenosis is associated with an exceptionally high risk of recurrent stroke. The outcomes of a prospective cohort investigation carried out in Hong Kong, China, among patients with ischemic stroke, revealed that individuals with intracranial or extracranial arterial stenosis who received intensified lipid-lowering therapy experienced a notably greater decline in the likelihood of recurrent ischemic stroke as opposed to their counterparts without such stenosis^27–29^. Statin and PCSK9i combination treatment stabilized intracranial atherosclerotic plaques more often compared to statins alone, as documented by HR-MRI^30^. The PCSK9i group showed a significant reduction in the stenosis degree. Our findings are in line with previous studies and also found a more pronounced benefit in this subgroup.

In this study, we included not only patients with large-artery atherosclerosis but also the embolic stroke of undetermined source (ESUS) with supra cardiac atherosclerotic plaque or mild stenosis (< 50%). The TOAST classification used an arbitrary threshold of 50% atherosclerotic stenosis to define large-artery atherosclerosis, whereas patients with a lower degree of stenosis were categorized as of undetermined cause^31^. Recent evidence suggested a significant aetiological association between supracardiac and stroke^32, 33^: carotid plaques are more prevalent ipsilateral to the infarct than contralateral^34^; aortic arch atherosclerosis is a frequent finding in ESUS patients^35^. ASCO phenotyping (A: atherosclerosis; S: small-vessel disease; C: cardiac pathology; O: other causes) indicates that nearly 90% of stroke patients exhibit some degree of atherosclerosis. Therefore, it is crucial to implement a systematic approach to controlling atherosclerotic risk factors in all cases of IS^2^.

In addition to reducing blood lipids, statins have anti-inflammatory, immunomodulation, and antioxidant properties, improving endothelial function and increasing nitric oxide bioavailability^36^. Statins improve outcomes in IS patients of atherosclerotic origin and other types of IS^37^. The cornerstone status of statins in the secondary prevention of IS is still unshakable. Whether evolocumab can have anti-inflammatory, as well as additional favorable non-lipid-lowering effects, needs further study.

Compared to statins or ezetimibe, evolocumab is more expensive and not recommended as a first-line treatment. The 2018 cholesterol guidelines from the ACC/AHA^9^ and 2019 ESC/EAS guidelines for the management of dyslipidemia^10^ recommended the method of “gradual lipid reduction”: PCSK9 inhibitors can be used for secondary prevention in patients who have a persistently elevated LDL-C level while receiving a maximally tolerated statin dosage, and only after first trying ezetimibe. We think the cost-effectiveness of the PCSK9 inhibitor can be improved by identifying and treating very high-risk patients who will get more absolute benefits from the treatment^38^. The sooner blood lipids reach the recommended treatment target, the earlier the risk of recurrent events can be reduced. Our study showed that the use of evolocumab in the early stages of IS was not associated with an increased incidence of hemorrhage or AEs. Therefore, for patients with AIS with a high risk of ASCVD, a rapid and early use of PCSK9 inhibitor is necessary to mitigate the risk of cerebrovascular events.

There are several limitations to our study. First, although we use propensity scores to control for confounding factors, many confounding factors such as baseline lipids and vascular stenosis cannot be fully controlled. Second, a retrospective study design can introduce several types of bias and limitations, including recall bias, selection bias, and incompleteness of data collection. Third, it was a single-center study design with a limited sample size. Our study included only urban individuals living in a city in China, limiting our results’ applicability to the whole of Chinese patients or other ethnicities and regions. These limitations could have led to overestimated effects of evolocumab on LDL-C-lowering. Based on the present results, multicenter prospective studies should be investigated further.

## Conclusions

This real-world study suggested that evolocumab on a background of statin reduced the LDL-C levels significantly with a well-tolerated profile and lowered the incidence of recurrent cerebrovascular events in the very high-risk ASCVD patients with AIS in China. However, further large real-world multicenter prospective studies or RCTs with long-term follow-up are warranted.

## Data Availability

We confirm that all data referenced in this manuscript will be made available upon request to qualified researchers for the purpose of replication and verification. The data will be available in a form that is consistent with ethical and legal requirements for data sharing and subject to any necessary institutional or ethical approvals.

## Acknowledgments

We express our gratitude to Dr. Yan Xu of the Department of Interventional Therapy, Tianjin Medical University Cancer Institute and Hospital, whose invaluable assistance in statistical analysis has greatly enriched our research endeavors.

## Author contributions

Conception and organization of the study: Ting Zhang, Yajing Zhang and Wei Yue.

Design and execution of the statistical analysis: Yajing Zhang and Ting Zhang.

Preparing the first draft: Ting Zhang.

Data collection and analysis, revision of the manuscript for intellectual content: Yun Yang, Haibing Liao, Xun Li, Ran Liu Xueqing Liu and Liqin Yang.

All authors have approved the final manuscript.

## Supplemental Material

Tables S1-S5 Figure S1-S6

